# Out-of-hospital Cardiac Arrest Before and During the COVID-19 Pandemic in the South Bronx

**DOI:** 10.1101/2022.06.08.22276169

**Authors:** Patrick Biskupski, Sebastian Ocrospoma Heraud, Mahmoud Khalil, Ali Horoub, Muhammad Maqsood, Kenneth Ong, Nail Cemalovic, Vidya Menon

**Affiliations:** Department of Internal Medicine, Lincoln Medical Center, New York City, NY, USA

## Abstract

**Background:** Out-of-hospital cardiac arrest (OHCA) is a major health challenge; the impact of the COVID-19 pandemic on OHCA in the South Bronx is unknown. The aim of this study was to determine differences between return of spontaneous circulation(ROSC), witnessed arrest, bystander CPR and survival to discharge, prior to and during the COVID-19 pandemic to improve ROSC and survival.

**Methods:** Single-center retrospective study of non-traumatic OHCA adult patients admitted to Lincoln Medical Center between 8/2019 to 6/2021, 3/2020 being the first established date of the COVID-19 pandemic in New York City. International Classification of Diseases (ICD) coding was used to identify cardiac arrests and collect information. Statistical analysis was performed using IBM-SPSS.

**Results:** ROSC time pre COVID-19 was 26 minutes, during the COVID-19 pandemic it was 25 minutes 54 seconds. A significant difference in witnessed arrests in the pre COVID-19 period compared to the COVID-19 period (86% vs 55% p = 0.03). Bystander CPR occurred 36% of the time in the pre COVID-19 period contrasting to 19% during. Prior to the COVID-19 pandemic the overall survival to discharge in OHCA ROSC cases was 28.5% comparing to 29% during the pandemic. ROSC was 18 minutes among survivors during the pandemic, compared to 21 minutes in survivors prior to COVID (p = 0.2).

**Conclusion:** There was a non-significant difference in ROSC, bystander CPR and survival to discharge in non-traumatic OHCA prior to and during the COVID-19 pandemic in the South Bronx. There was a significant difference in witnessed vs unwitnessed OHCA prior to and during the COVID-19 pandemic.

## Introduction

Out-of-hospital cardiac arrest (OHCA) represents loss of cardiac function in the outpatient setting. Cardiovascular collapse interrupts global perfusion with subsequent cell death. Cardiopulmonary resuscitation (CPR) is an emergency lifesaving procedure performed by the general public or health care professionals in attempts to liberate systemic perfusion and eventual resuscitation of spontaneous circulation (ROSC). Despite global initiatives to educate communities on performing and understanding algorithms involving CPR, OHCA remains a major health challenge. The Cardiac Arrest Registry to Enhance Survival (CARES) is a national registry of OHCA with data collected from numerous districts, providing communities with outcomes used to develop and implement strategies to increase ROSC rates and improve survival to discharge [1]. However, CARES and registry’s alike do not represent the underserved communities like South Bronx. With the onset of the COVID-19 pandemic, the impact of the pandemic on OHCA in the South Bronx is unknown. The aim of our study was to determine differences between return of spontaneous circulation(ROSC), witnessed vs unwitnessed arrest, bystander CPR, patient characteristics and survival to discharge, prior to and during the COVID-19 pandemic in the South Bronx.

## Materials & Methods

The Bronx is one of the five boroughs of New York City, USA. This study was performed at Lincoln Medical Center a community teaching hospital with 362 beds located in the heart of the South Bronx, with approximately 164,979 emergency department visits annually the third highest nation-wide. The South Bronx with a population of 522 412 represents 40% of the total population of the Bronx with 40% of its inhabitants below the federal poverty line [2].

This was a single-center retrospective study of all non-traumatic OHCA adult (18 years or older) patients admitted to LMC between 8/2019 to 6/2021, with 3/2020 being the first established date of the COVID pandemic in New York City. International Classification of Diseases (ICD) coding was used to identify OHCA, and information was collected pertaining to demographics, comorbidities, laboratory results, treatments and outcomes and entered into a centralized database. Traumatic OHCA, pediatric and inpatient cardiac arrests were excluded.

Statistical analysis was performed using IBM Corp. 2021. IBM SPSS statistics for Macintosh, version 26.0. Specifically, descriptive statistics and nonparametric tests. Or study was approved by the institutional review board (IRB) at Lincoln Medical Center (LMC).

## Results

Figure 1 displays the exclusion criteria for patients in our study. A total of 535 patients with cardiac arrest were admitted to Lincoln Medical Center (LMC) between 8/2019 and 6/2021. After exclusion of traumatic and inpatient arrests the total became 114 OHCA sustaining ROSC. Of the total, 28 cases were admitted pre COVID-19 (8/2019 to 6/2021) of which 8 patients survived to discharge. 86 cases admitted during the COVID-19 pandemic (3/2020-6/2021) with 25 patients surviving to discharge.

**Figure 1.**
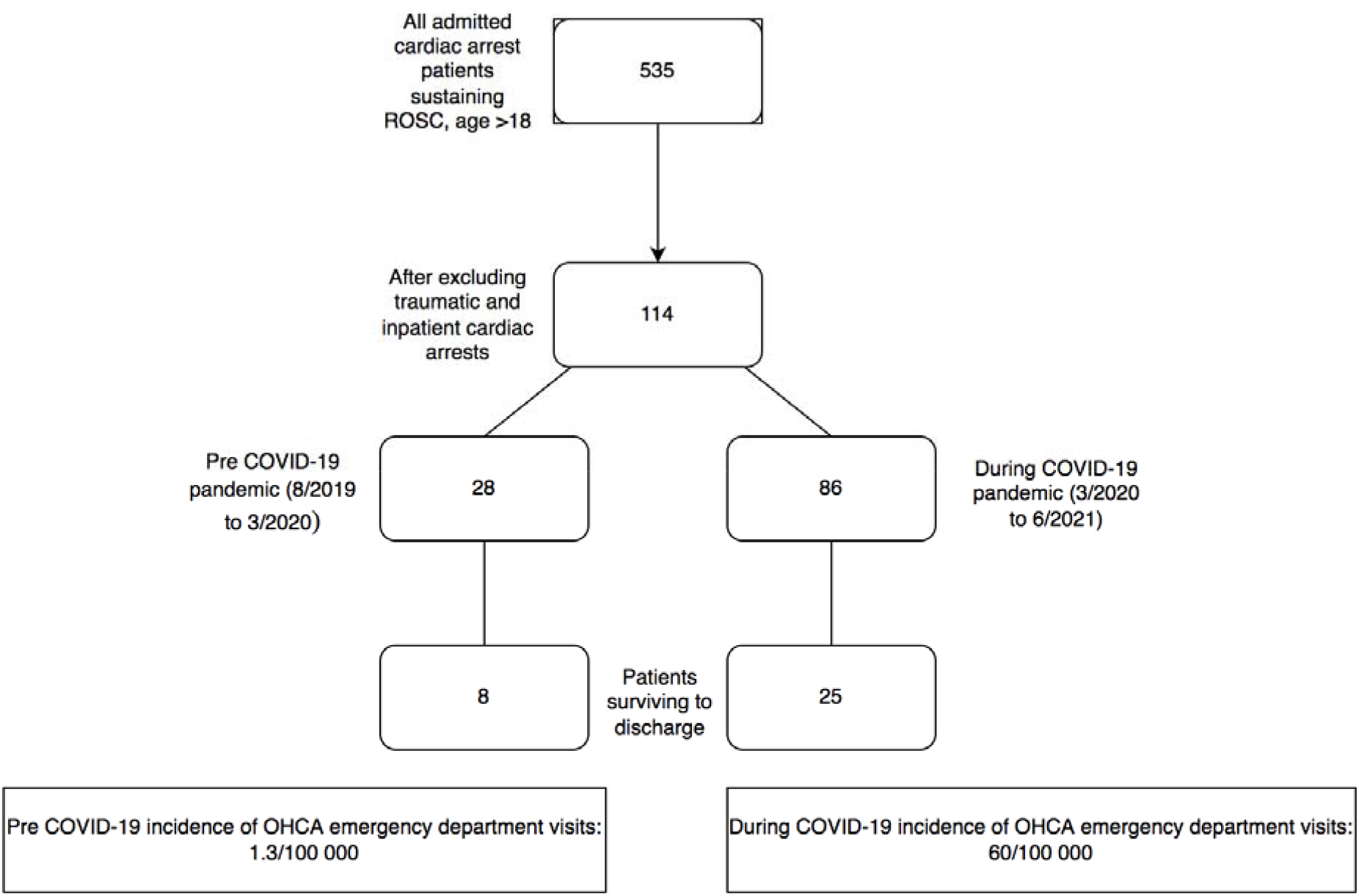

Table 1 illustrates patient characteristics during OHCA sustaining ROSC irrespective of survival to discharge during the pre-pandemic and pandemic period. The mean ROSC time pre COVID-19 was 26 minutes, during the COVID-19 pandemic it was 25 minutes 54 seconds, non statistically significant (p-value: 0.942). The location of cardiac arrest was predominantly at home in both groups, with a significant association of witnessed arrests in the pre COVID-19 period compared to the COVID-19 period (86% vs 55% p = 0.03). The first documented rhythm was frequently non shockable, with bystander CPR occurring 36% of the time in the pre COVID-19 period contrasting to 19% during. Baseline patient characteristics including age, gender were comparable in our predominantly Latin population. Comorbidities of patients in both cohorts were comparable except for a significantly higher number of patients during the COVID period with asthma (19% vs 1% p = 0.004) and extreme obesity (22% vs 11%). TTM was comparable.

**Table 1.**
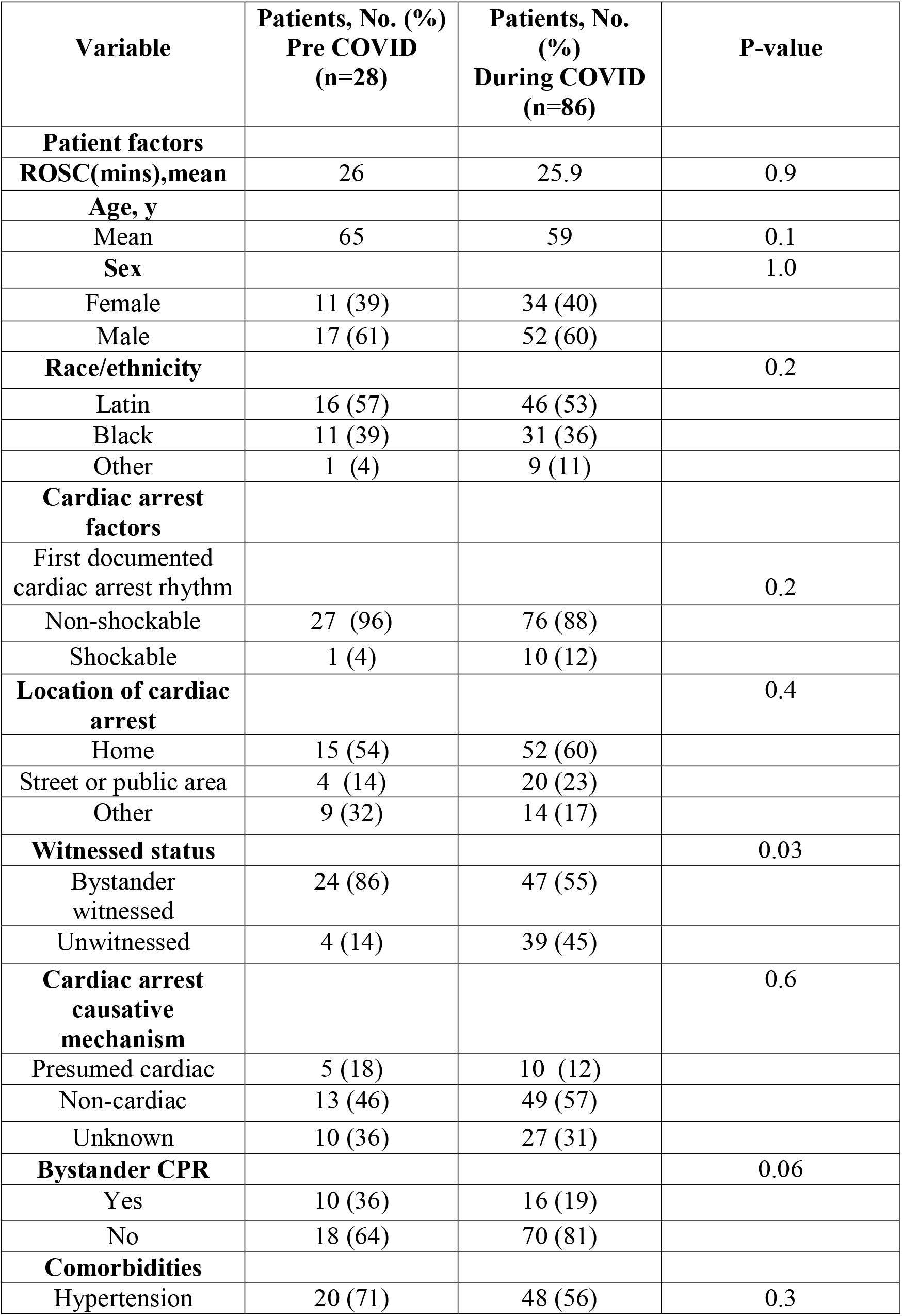
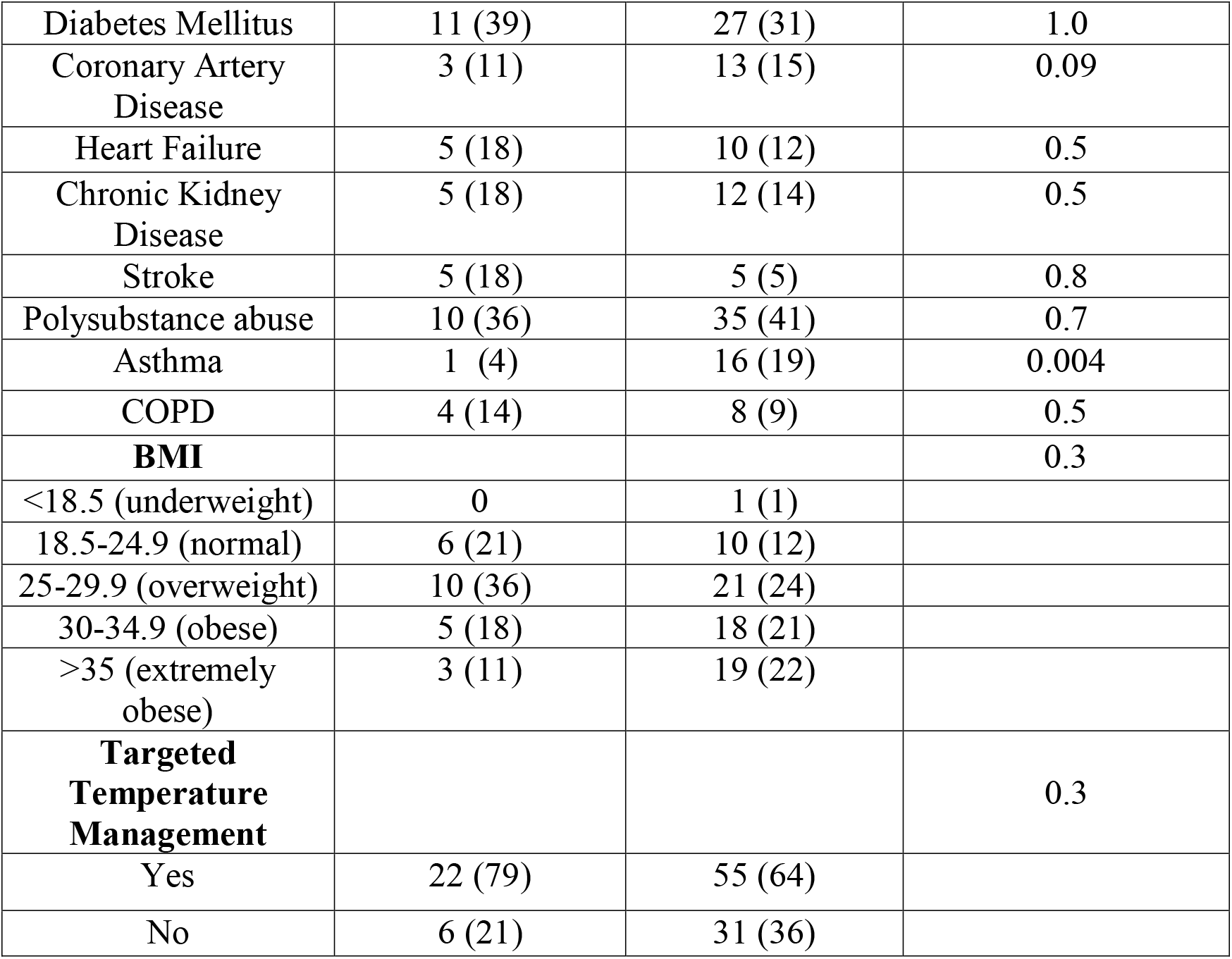
Characteristics of patients prior to the COVID-19 pandemic (8/2019 - 3/2020) and during (3/2020 - 6/2021)

Table 2 demonstrates characteristics of OHCA ROSC cases surviving to discharge in the pre-pandemic (8/2019 - 3/2020) and during (3/2020 - 6/2021). Prior to the COVID-19 pandemic the overall survival to discharge in OHCA ROSC cases was 28.5% comparing to 29% during the pandemic. Contrasting with only 9% of OHCA surviving to hospital discharge per 2020 OHCA CARES(Cardiac Arrest Registry to Enhance Survival) summary reportc[1]. Baseline characteristics of survivors in both groups were comparable including age, gender and ethnicity. ROSC was 18 minutes among survivors during the pandemic, compared to 21 minutes in survivors prior to COVID (p = 0.2). While the majority of the survivors in both groups had cardiac arrest at home, 88% of the pre-COVID-19 survivors had witnessed arrest compared to 60% during the COVID-19 pandemic. Bystander witnessed cardiac arrest among survivors of OHCA as per the 2020 CARES summary report was only 37% [1]. Survival to discharge among patients with bystander-initiated CPR was 25% prior to, compared to 12% during the pandemic. This contrasts with 41% survival among patients with OHCA when bystander-initiated CPR as per the 2020 CARES summary report [1]. The comorbidities among the survivors in the pre-COVID-19 period and during the COVID-19 pandemic were comparable, with 50% of survivors to discharge having a history of polysubstance use. All the survivors in the pre-COVID-19 and 86% of survivors in during the COVID-19 period received TTM. While majority (50-60%) of the survivors were discharged to skilled nursing facility (SNF), 12% of pre-COVID-19 and 8% of COVID-19 period survivors required a long term acute care hospital. No survivors were discharged home.

**Table 2.** Characteristics of patient’s surviving to discharge prior to the COVID-19 pandemic (8/2019 - 3/2020) and during (3/2020 - 6/2021)

| **Variable** | **Patients, No. (%)**<br>**Pre COVID**<br>**(n=8)** | **Patients, No. (%)**<br>**During COVID**<br>**(n=25)** | **P-value** |
| --- | --- | --- | --- |
| **Patient factors** |  |  |  |
| **ROSC(mins),mean** | 21 | 18 | 0.2 |
| **Age(y), mean** | 65 | 54 | 0.06 |
| **Sex** |  |  | 0.07 |
| Female | 2(25) | 10 (40) |  |
| Male | 6(75) | 15 (60) |  |
| **Race/ethnicity** |  |  | 1.0 |
| Latin | 6(75) | 10 (40) |  |
| Black | 2(25) | 11 (44) |  |
| Other | 0 | 4 (16) |  |
| **Cardiac arrest factors** |  |  |  |
| First documented cardiac arrest rhythm |  |  | 0.3 |
| Non-shockable | 8(100) | 19 (76) |  |
| Shockable | 0 | 6 (24) |  |
| **Location of cardiac arrest** |  |  | 0.3 |
| Home | 4(50) | 12 (48) |  |
| Street or public area | 3(38) | 8 (29) |  |
| Other | 1(12) | 5 (23) |  |
| **Witnessed status** |  |  | 0.2 |
| Bystander witnessed | 7(88) | 15 (60) |  |
| Unwitnessed | 1(12) | 10 (40) |  |
| **Cardiac arrest causative mechanism** |  |  | 0.4 |
| Presumed cardiac | 0 | 4 (16) |  |
| Non-cardiac | 5(63) | 17 (68) |  |
| Unknown | 3(7) | 4 (16) |  |
| **Bystander CPR** |  |  | 0.6 |
| Yes | 2(25) | 3 (12) |  |
| No | 6(75) | 22 (88) |  |
| **Comorbidities** |  |  |  |
| Hypertension | 5(63) | 12 (48) | 0.4 |
| Diabetes Mellitus | 3(38) | 4 (16) | 0.7 |
| Coronary Artery Disease | 0 | 2 (8) | 0.4 |
| Heart Failure | 1(13) | 4 (16) | 0.9 |
| Chronic Kidney Disease | 1(13) | 3 (12) | 0.7 |
| Stroke | 1(13) | 0 | 1 |
| Polysubstance abuse | 4(50) | 13 (52) | 0.2 |
| Asthma | 0 | 6 (21) | 0.2 |
| COPD | 0 | 1 (4) | 0.4 |
| **BMI** |  |  | 0.1 |
| <18.5 (underweight) | 0 | 0 |  |
| 18.5-24.9 (normal) | 3 | 3 (12) |  |
| 25-29.9 (overweight) | 2 | 7 (28) |  |
| 30-34.9 (obese) | 3 | 8 (32) |  |
| >35 (extremely obese) | 0 | 2 (8) |  |
| **Targeted Temperature Management** |  |  | 0.6 |
| Yes | 8(100) | 21 (84) |  |
| No | 0 | 4 (16) |  |
| Length of Stay, days, mean | 37.9 | 23.1 | 0.2 |
| **Disposition** |  |  | 0.1 |
| Home | 0 | 0 |  |
| SNF | 4(50) | 15(60) |  |
| LTACH | 1(12.5) | 2(8) |  |
| Hospice | 2(25) | 0 |  |

## Discussion

CARES is a national registry of OHCA with data collected from numerous districts, providing communities with outcomes used to develop and implement strategies to increase ROSC rates and improve survival to discharge. However, CARES and registry’s alike do not represent the underserved community of the South Bronx. Our study is the first such study of OHCA in the South Bronx. Incidence of OHCA in the South Bronx prior to the pandemic (8/2019 – 3/2020) was 1.3/100,000 emergency department (ED) visits, during the pandemic (3/2020 – 6/2021) it increased to 60/100,000 ED visits in our community. This trend was evident globally (United Kingdom, Holland, Spain, Sweden, Australia, France, Italy, Korea, Singapore) [3]. We reported similarities between OHCA ROSC time, bystander CPR, non-shockable rhythms during both time periods. Similar rates of bystander CPR during both periods were noted by Lai et al [4]. No changes in ROSC rates were observed with OHCA in Detroit by a study conducted by Matthew et al [5]. Contrasting to studies exhibiting decrease in ROSC during the pandemic [6]. It can be hypothesized that the pandemic had a greater impact in communities with baseline favorable outcomes in ROSC, less non shockable arrests, tendency towards bystander CPR and less comorbidities in comparison to underserved communities like the South Bronx.

Survival to discharge rates vary amongst studies in other communities and the CARES data summary report [1]. In our study prior to the pandemic the overall survival to discharge in OHCA ROSC cases was 28.5% versus 29% during the pandemic, similarities also evident by Semeraro et al [7]. Contrasting to the majority of studies reporting a lower percentage of survival to hospital discharge during the pandemic relative to the pre-pandemic period. Moreover, only 9% of OHCA surviving to hospital discharge per 2020 OHCA CARES summary report [1]. We observed a large portion of non-shockable arrests pre and during the pandemic, signifying that the majority of OHCA are likely not of cardiac origin [8;9;10]. It must be noticed that 50% of survivors’ pre-pandemic and 52% of survivors during the pandemic had a history of polysubstance abuse, unique to our South Bronx population. Half of the neighborhood tabulation areas in NYC with the highest drug related hospitalization are in the Bronx [11]. Drug overdose a possible cause of non-shockable cardiac arrests in our population, with a high propensity occurring at home with a greater chance of survival among them correlated with our results and a study by Orkin et al [12]. We observed that none of our survivors were discharged home.

Although survival to discharge in our population was higher per national standards [1]. Bystander -initiated CPR (cardiopulmonary resuscitation) was scarce 42% pre-pandemic vs 34% in the COVID-19 period. Despite a predominant percentage of cardiac arrests being witnessed (86% pre-pandemic vs 55% during pandemic) and located at home (54% pre-pandemic vs 60% during). Bystander CPR increases survival of OHCA by 2 to 3 times [13]. Increasing willingness of bystanders to perform CPR has been a long-term challenge in the pre-pandemic age, with the general public’s fear of injury, lack of skill and knowledge. With the introduction of the pandemics new limitations bystander CPR rates have diminished further, compounded by fear of contagion. This was highlighted by surveys on the general public’s inclination to provide CPR after witnessing a OHCA, with an unexpected decrease in CPR to strangers by HCW (healthcare workers) comparing to non HCW, improving with PPE availability [14]. In our community reinstating CPR training in schools and work places, with increased availability of emergency PPE, could improve the rates of bystander CPR. We believe that our results can be used to improve OHCA and survival to discharge in our community and be expanded to other underserved communities during the COVID-19 era.

## Limitations

Our study had several limitations. Firstly, emergency medical service (EMS) variables were not included in this study including time intervals (departure and arrival), chest compression interruptions and airway difficulties. Secondly, we did not collect any information on COVID-19 status in the admitted patients nor the vaccination status. This could have accounted for the majority of OHCA during the period of the pandemic in our cohort. As COVID-19 is known to induce hypoxic respiratory failure, pulmonary emboli and infarction ultimately resulting in cardiac arrest with a non-shockable rhythm [15]. Furthermore, cerebral performance data was not available at discharge, preventing prognostication of long-term survival, neurological and functional status. It has been shown that cerebral performance can predict long term survival outcomes of OHCA [16]. Finally, as this study was performed at the start of the COVID-19 pandemic, it is unknown if the ROSC and survival has since changed with the introduction of the vaccine. Lastly, this study was conducted at a single center, limiting applicability of our results to other settings.

## Conclusions

The purpose of this study was to determine differences between return of spontaneous circulation(ROSC), witnessed arrest, bystander CPR and survival to discharge, prior to and during the COVID-19 pandemic to improve ROSC and survival. Our results show a non-significant difference in ROSC, survival to discharge, bystander CPR and patient characteristics in non-traumatic OHCA admitted to Lincoln Medical Center prior to and during the COVID-19 pandemic in the South Bronx. There was a significant difference in witnessed vs unwitnessed OHCA prior to and during the COVID-19 pandemic. We believe that our results can be used to improve OHCA and survival to discharge in our community and be expanded to other underserved communities during the COVID-19 era.

## Data Availability

Requests for de-identified data may be directed to the corresponding authors and will be reviewed by the research department at Lincoln Medical Center. Data limitations are designed to ensure patient confidentiality.

## Acknowledgements

Not applicable

